# Uncovering and Understanding Success: A Qualitative Study of High-Performing Hospitals for Small and Sick Newborn Care in Four countries in Africa

**DOI:** 10.1101/2025.11.12.25339979

**Authors:** Kylie Dougherty, Ifeanyichukwu Anthony Ogueji, Hannah Mwaniki, Jitihada Baraka, Samuel Ngwala, Lucky Mangwiro, Lucas Malla, Positive outlier facility staff group authorship, Mariam Johari, Chinyere Ezeaka, Kondwani Kawaza, Robert Tillya, Donat Shamba, Z. Maria Oden, Rebecca Richards-Kortum, Natasha Rhoda, Joy E. Lawn, Lisa R. Hirschhorn, Christine A. Bohne

**Affiliations:** Department of Medical Social Sciences and Havey Institute for Global Health, Feinberg School of Medicine, Northwestern University, Chicago, IL, USA; APIN Public Health Initiatives, Nigeria; Aga Khan University, Nairobi, Kenya; Ifakara Health Institute, Department of Health Systems Impact Evaluation and Policy, Dar es Salaam, Tanzania; Kamuzu University of Health Sciences, Malawi, NEST360 Project; London School of Hygiene and Tropical Medicine, London, UK; University of Lagos, College of Medicine, Lagos, Nigeria; Rice University, Rice360 Institute for Global Health Technologies, Houston, USA; University of Cape Town, Cape Town, South Africa

**Keywords:** Positive outliers, quality improvement (QI), small and sick newborn care, sub-Saharan Africa

## Abstract

**Background:** Despite evidence-based interventions to reduce neonatal mortality, implementation gaps continue in low-resource settings. The Newborn Essential Solutions and Technologies (NEST360) alliance supports neonatal units in Kenya, Malawi, Nigeria, and Tanzania with a package of technologies, training, data systems, and quality improvement (QI). However, wide variation in care coverage remains. This study examines the facility-level strategies used by high-performing “positive coverage outlier” hospitals to achieve and sustain high coverage of priority neonatal interventions.

**Methods:** We performed 74 key informant interviews and 21 focus groups across 16 hospitals in four countries. Hospitals were selected based on their coverage rates (percentage of eligible newborns receiving an intervention) for at least one of four neonatal interventions (kangaroo mother care, continuous positive airway pressure, phototherapy, hypothermia prevention). Transcripts were coded using deductive and inductive approaches to identify strategies, barriers, and facilitators of implementation.

**Results:** A total of 114 distinct strategies were identified. Common strategies across countries included peer-to-peer mentorship, routine data review meetings, and family education. Country-specific adaptations were also observed. For example, Malawi emphasized data-driven decision-making and Tanzania focused on infection control. Many strategies were low-cost and addressed organizational and behavioral mechanisms such as motivation, accountability, and teamwork.

**Conclusions:** This study highlights the importance of context-sensitive implementation strategies in achieving and sustaining high coverage for neonatal interventions. Beyond program interventions, like trainings and providing medical devices, strategies fostering staff engagement, leadership support, and cultural alignment are critical. Findings offer actionable insights for scaling up neonatal care improvements within and beyond the NEST360 network.

**Clinical trial number:** Not applicable

**Contributions to the literature:**

1. This study demonstrates how implementation science principles can be applied to facility-level quality improvement efforts to generate generalizable knowledge and identify common strategies and themes that underpin successful implementation across diverse facilities and countries.
2. These findings highlight the variability in program implementation across facilities and countries, demonstrating that even when facilities receive the same package, success depends on context-specific adaption. This contributes to the growing evidence that effective scale-up requires flexible, locally tailored solutions in additional to a standardized implementation package.
3. The use of a positive outlier approach within a multi-country implementation program demonstrates a feasible and replicable method for identifying and disseminating actionable implementation strategies, and the cross-country comparative design of this study enables the identification of convergent and context-specific strategies used to improve small and sick newborn care.

## 1. Background

Newborn mortality remains a significant global health challenge, particularly in low-resource settings where the burden is highest (1). While numerous evidence-based interventions (EBIs) have been developed and identified to improve neonatal health outcomes, there is still a gap between evidence and practice (2). For example, interventions such as kangaroo mother care (KMC), which promotes skin-to-skin contact and breastfeeding, and continuous positive airway pressure (CPAP) therapy for managing neonatal respiratory distress, are proven to reduce morbidity and mortality (3, 4). However, their success hinges not only on clinical effectiveness but also on the extent to which these interventions are delivered consistently and with fidelity within the constraints of the local health system (5). Factors such as provider training, supportive supervision, caregiver engagement, policy support, and functional referral systems all influence whether these interventions translate into meaningful impact on the ground (6).

The Newborn Essential Solutions and Technologies (NEST360) alliance addresses the implementation gap through a comprehensive, multi-country implementation program aimed at improving care for small and sick newborns (7). NEST360 supports hospitals in Kenya, Malawi, Nigeria, Ethiopia, and Tanzania that have a level 2 or higher neonatal unit. The world health organization defined level 2 neonatal care units as those that provide care for moderately ill and extremely ill newborns, including those with hemodynamic instability, respiratory distress requiring CPAP or short-term ventilation, very low birth weight, abnormal neurological exams, seizures, or those needing exchange transfusions (8). The program deploys a coordinated package that includes technology bundles, clinical and biomedical training, data systems, and facility-level quality improvement (QI) support (7).

However, despite receiving the same core interventions, NEST360 hospitals exhibit wide variation in both care coverage and quality for key interventions—including KMC, CPAP, hypothermia prevention, and phototherapy. These variations in intervention coverage help to explain the variation in mortality reduction that NEST360 found in their interim analysis (9). For example, among NEST360-supported hospitals, some hospitals had not seen a significant mortality reduction, while others saw a 54% reduction (9). This variation highlights the critical importance of identifying context-specific solutions used by the higher-performing hospitals. This same pattern of wide variations in program outcomes despite standardized interventions has been observed in other global health programs, reinforcing the need to understand the drivers of care variations and identify context-specific solutions to remedy them (10–12).

These contextual solutions, which are activities that are tailored to work in specific settings based on the realities of that context, are actions that drive change and are often implemented through QI initiatives, which use systematic, data-guided identification of barriers to design and test activities to bring about improvements in healthcare delivery (13, 14). Locally generated change ideas and iterative improvement projects have helped teams identify and address gaps in care processes, leading to measurable improvements in service delivery (15, 16). However, these facility-level efforts are often more contextualized and not disseminated widely, even if their solutions have potential benefit for other settings (17). This study aimed to explore the activities and change ideas across multiple NEST360-supported hospitals in four countries (Kenya, Malawi, Nigeria, Tanzania) that either improved or sustained high coverage rates of priority neonatal interventions (eg, KMC, CPAP, hypothermia prevention, phototherapy), which is the percentage of eligible newborns receiving a care intervention. The goal was to examine not only *what* worked in specific hospitals but also *how* and *why* certain strategies succeeded, with the ultimate goal of generating generalizable knowledge that can be shared with other hospitals and accelerate progress toward reducing neonatal mortality across diverse settings.

## 2. Methods

### Theoretical frameworks

We used the NEST360/United Nations Children’s Fund (UNICEF) small and sick newborn care (SSNC) 10 core components framework, as adapted by the Newborn Toolkit community, to identify and organize barriers, enablers, and strategies driving the variations in clinical coverage seen in the positive outliers’ performance (see Figure 1) (18, 19). This framework was selected because it focuses on the key components of the health system building blocks that impact SSNC delivery and EBI utilization (18, 19). We used the Standards for Reporting Qualitative Research (SRQR) (see Supplemental Document 1) to guide the reporting of this study.

**Figure 1.**
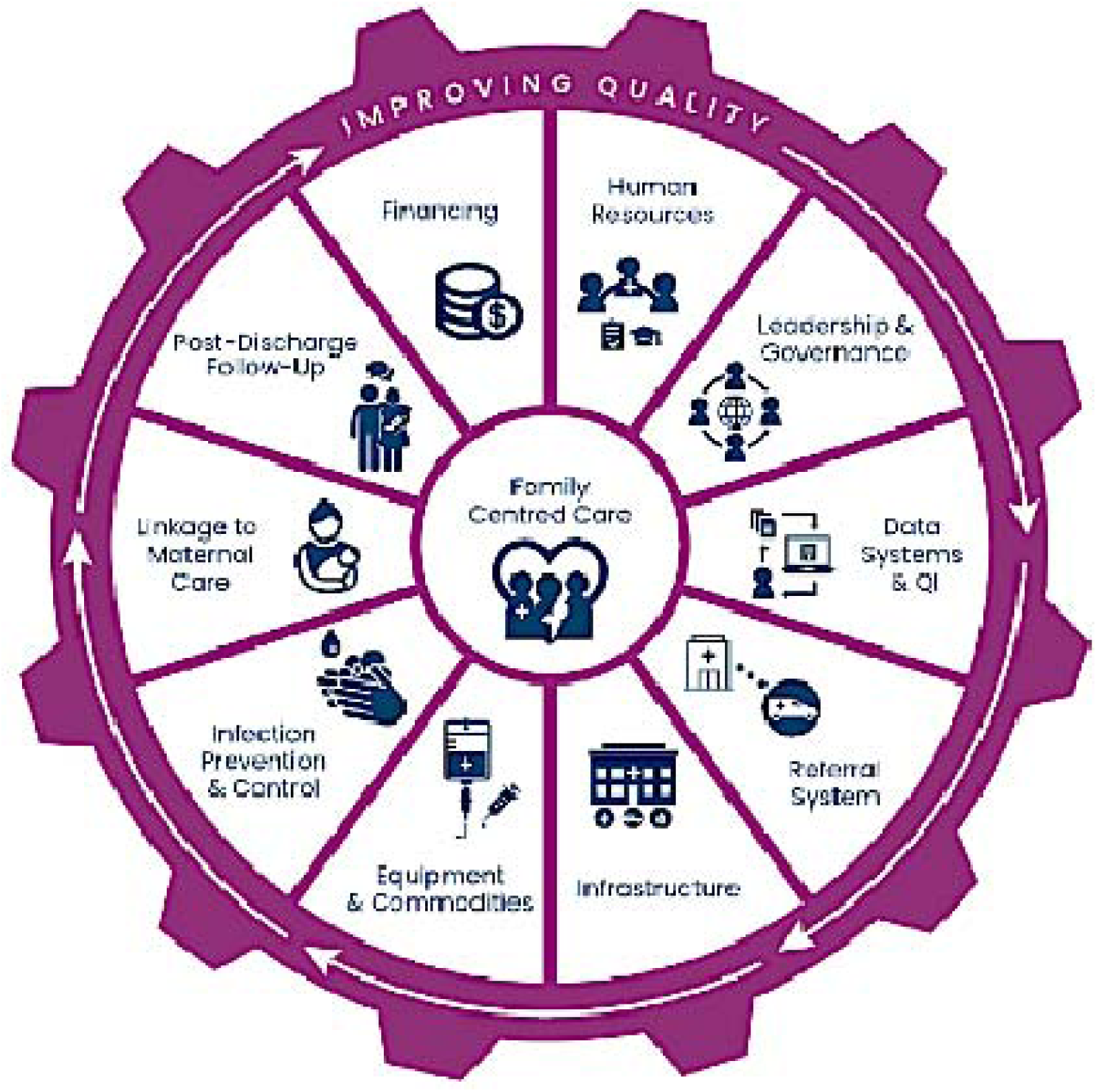
NEST360 and UNICEF’s Small and Sick Newborn 10 Core Components Framework(20)

### Sample Selection

This study examined data across 65 hospitals in four sub-Saharan African countries – Kenya, Malawi, Nigeria, and Tanzania. All participating facilities were Level 2+ neonatal units engaged in the implementation of NEST360’s priority interventions. An interim analysis carried out in the first quarter of 2023 indicated that four interventions – phototherapy for neonatal jaundice, KMC, hypothermia prevention, and CPAP – collectively explained approximately 98% of the observed variation in mortality change; and were therefore identified as the primary drivers of mortality reduction. In contrast, interventions such as antibiotic use for sepsis already had consistently high coverage, leaving limited scope for evaluating their causal contribution. Others, such as blood cultures for infection, had very low denominators since cultures are rarely performed in these settings. Consequently, our analysis focused on coverage changes in phototherapy, KMC, hypothermia prevention, and CPAP.

Coverage was defined as the percentage of eligible newborns who received a given intervention. For example, CPAP coverage referred to the proportion of newborns who received CPAP among all those eligible for it. The analysis examined changes in coverage between two six-month periods: January–June 2023 and July–December 2023. Coverage change was calculated as the difference in coverage between Q1–Q2 (Jan–Jun) and Q3–Q4 (Jul–Dec) of 2023. In cases where the number of eligible newborns (the coverage denominator) for an intervention in Q3–Q4 of 2023 was too small (fewer than 20), the analysis was extended to a 12-month comparison, using coverage data from 2022 versus 2023. The analysis used a total of 95,260 neonatal admission records across the 65 hospitals.

To explore the activities driving clinical coverage variation amongst NEST360-supported hospitals, we employed an outlier case selection approach. In each country, national NEST360 teams purposively identified a minimum of four hospitals considered positive outliers for at least one of four priority clinical indicators: CPAP, KMC, phototherapy, and hypothermia prevention. This study classified positive coverage outliers into two groups: sustainers, defined as hospitals maintaining at least 80% coverage across two consecutive six-month periods in 2023, and rapid progressors, defined as those in the top 25% for coverage improvement over the same periods (see Figure 2).

**Figure 2:**
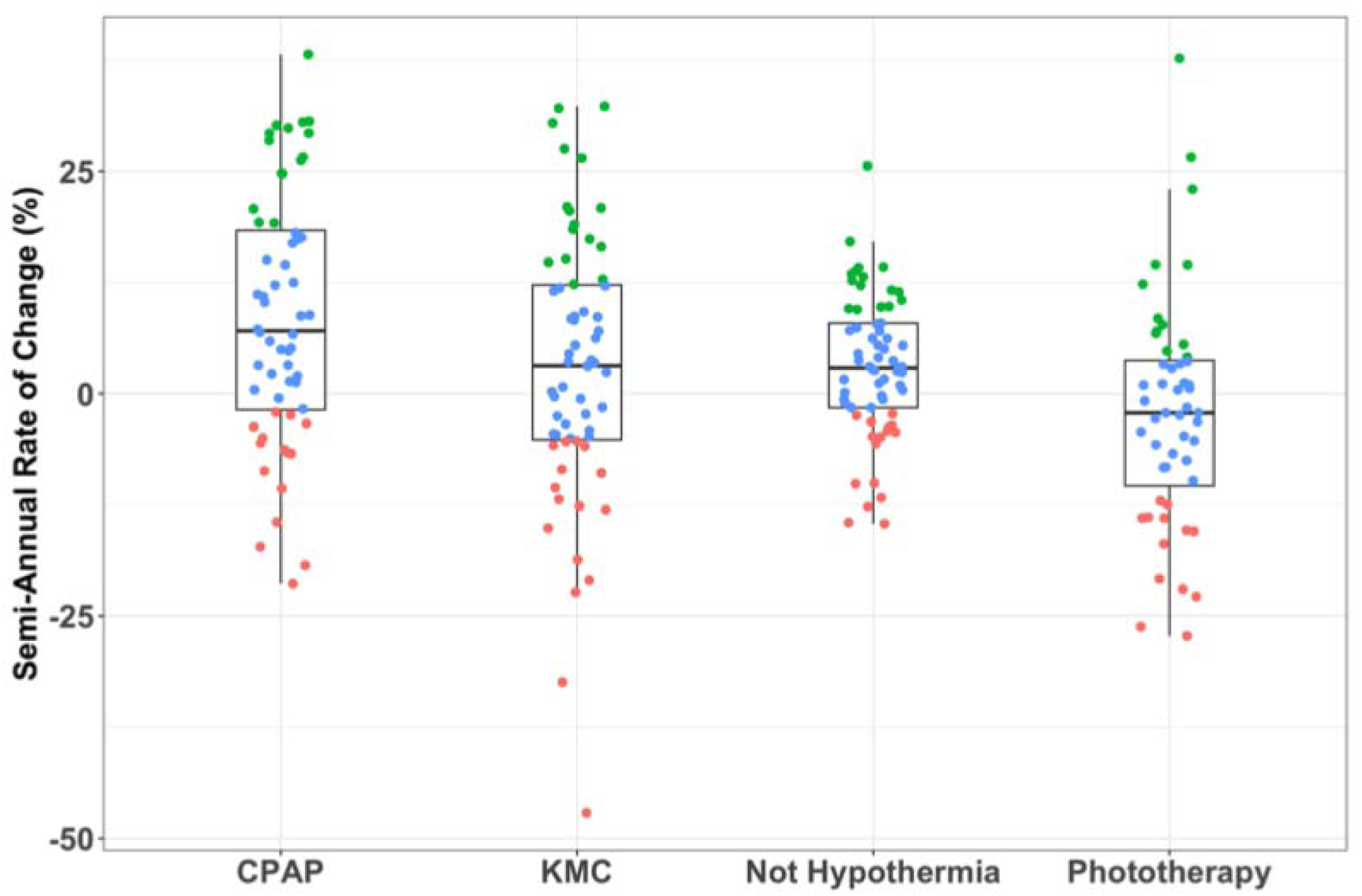
Coverage changes between Q1–Q2 (Jan–Jun) and Q3–Q4 (Jul–Dec) of 2023. *Each dot represents a facility – and green shows rapid progressors in top 25% of improvements*.

To choose the included sites, country team members were provided with a list of sites that met the criteria for sustainer or rapid progressor outlier to choose from. Additional sites based on the availability of outliers set by these categories were selected based on the country teams’ contextual knowledge of facility-level QI efforts. A total of 18 sustainer and 36 rapid progressor hospitals were identified. Figure 3 presents a flow diagram for the selected positive coverage outliers, and Supplemental Table 1 shows the clinical indicators that each positive coverage outlier met.

**Figure 3.**
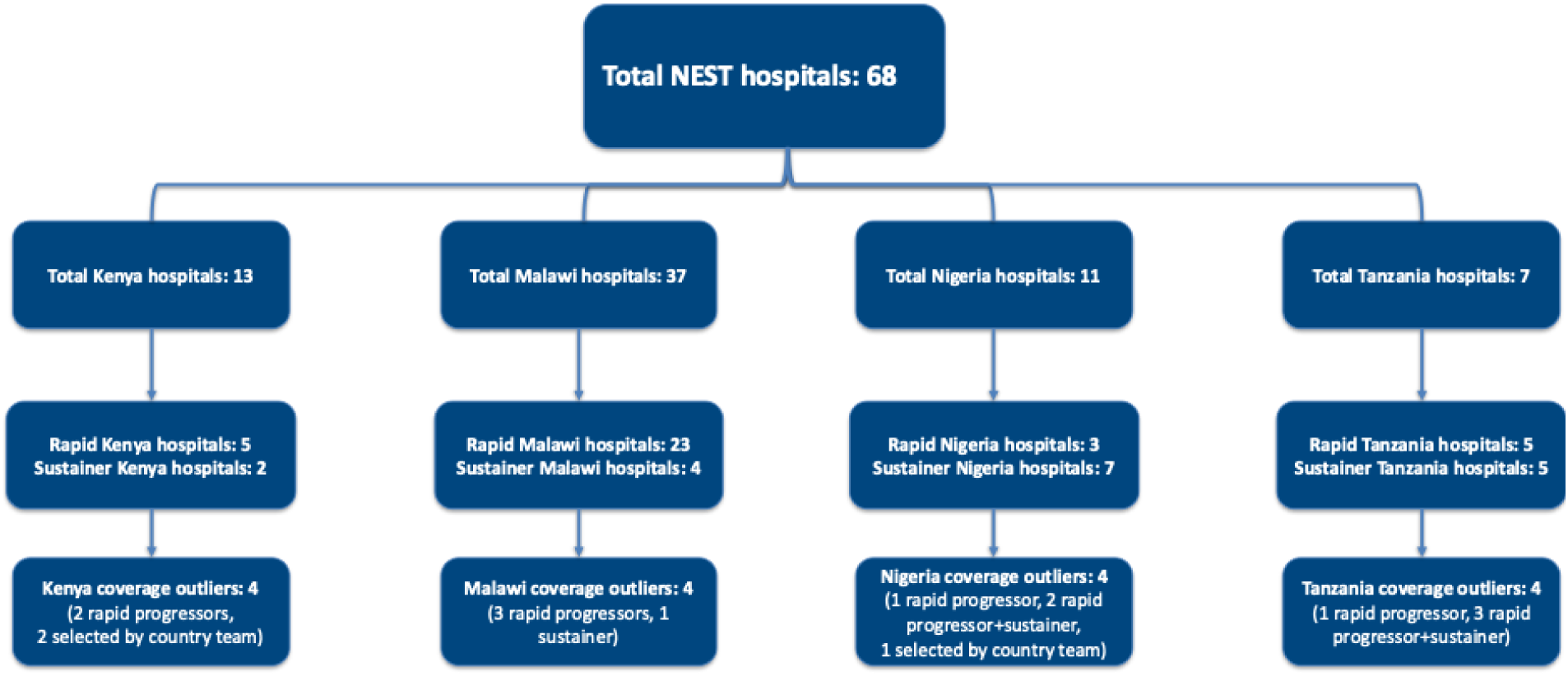
Flow chart of facilities selected to be positive coverage outliers *Note.* A hospital may be both a rapid progressor for some clinical indicators and a sustainer for others (e.g., rapid progressor in KMC and CPAP, but sustainer in hypothermia prevention). Thus, the number of outlier hospitals does not equal the total number of hospitals.

Participants were purposively sampled from the selected outlier hospitals and included a range of clinical and support staff engaged in neonatal care and improvement initiatives. These included QI leads, biomedical engineering technicians (BMETs), data clerks, nurses, nurse leaders, physicians, and members of hospital management. This diverse group of respondents provided insight into facility-specific strategies and contextual factors influencing implementation and care coverage.

### Data Collection

Data collection involved key informant interviews (KIIs) and Focus Group Discussions (FGDs), conducted in person at hospitals. A NEST360-trained qualitative researcher led the data collection process in each of their countries, ensuring consistency and methodological rigor. These qualitative researchers had received qualitative training from NEST360 and completed previous studies for NEST360 (21). The interview guide focused on exploring barriers and facilitators that hindered or supported the provision of SSNC, focusing on the four priority EBIs, as well as actions taken and QI project activities that participants believed contributed to improved or sustained high coverage of key indicators. The guide was iteratively refined with input from the research team to enhance relevance and clarity. Interviews and focus groups were conducted by a native speaker in Swahili (Tanzania), Chichewa (Malawi), English (Nigeria), and English (Kenya). All participants provided verbal consent prior to participating in the study, including consent to be audio recorded. Data were captured through audio recordings and field notes, ensuring comprehensive documentation of insights. Each data collection session lasted between 30 to 60 minutes, balancing depth of discussions with participant engagement. Member checks were performed during the interviews, as data collectors summarized key concepts and ideas that the participants voiced and asked for confirmation to ensure perspectives were being accurately captured.

### Data Management and Preparation

Audio files were stored on OneDrive (Microsoft Corporation), an encrypted, password-protected electronic site accessible only to a research team member (22). All participants were identified by a unique participant number. All audio files were transcribed and blinded by the NEST360 team member who conducted the KIIs and FGDs and were checked for accuracy. All audio files not in English were translated by the data collector, who was fluent in both the local language and English.

### Data Analysis

A rigorous qualitative descriptive methodology approach was applied across four countries to ensure reliability, validity, and contextual depth (23). A dual-coding strategy was used for each country, with at least two coders involved: KD and the respective country lead (HM, SN, LM, JB, IAO). KD served as the only coder to analyze transcripts across all countries, ensuring consistency in interpretation. The inclusion of country-level qualitative researchers ensured a contextual understanding of the environment and supported an in-depth analysis of the findings.

The coding process combined deductive and inductive approaches using NVivo software. Deductive coding was structured around predefined domains and parent codes (see codebook in Supplemental Document 2), while inductive coding allowed for the emergence of sub-codes related to facility and country-specific strategies, barriers, and facilitators. The preliminary codebook was developed a priori, with KD developing the initial version and then having HM, SN, LM, JB, and IAO revise it. The coding framework for the codebook was organized based on the SSNC core components (20), such as *data systems* or *human resources*. Subcodes were developed iteratively throughout the coding process as the coders met weekly to discuss emerging findings. Emerging themes surrounding barriers, facilitators, and strategies were identified during the coding process, reflecting both pre-existing concepts and newly uncovered insights through transcript analysis.

### Ethics

Ethical approval for this study was conducted by London School of Hygiene and Tropical Medicine IRB (21892), and the national health research ethics committee in Malawi (NHSRC 1180), Kenya (Kenya Medical Research Institute: KEMI/SERU/CGMR-C/229/4203, Maseno University Ethics Review Committee: MSU/DRPI/MUERC/00810/19), Tanzania (Ifakara Health Institute: IHI/IRB/No:01-2020, National Institute for Medical Research: NIMR/HQ/R.8a/Vol.IX/3405), and Nigeria (Lagos University Teaching Hospital Health Research Ethics Committee: ADM/DCST/HREC/APP/3487, University of Ibadan/University College Hospital/Ethics Committee: NHREC/05/01/2008a).. The study was conducted with the ethical principles outlined in the Declaration of Helsinki. All participants were informed about the purpose of the study, assured of confidentiality, and provided verbal consent prior to participation.

Participation was voluntary, and respondents could withdraw at any time without consequence.

## 3. Results Participant demographics

One hundred and thirty-two participants were recruited into this study. Overall, we performed 21 FGDs and 74 KIIs. Table 1 presents a breakdown of participants by country and job type. The participants identified 109 strategies that they believed influenced their high performance.

**Table 1.**
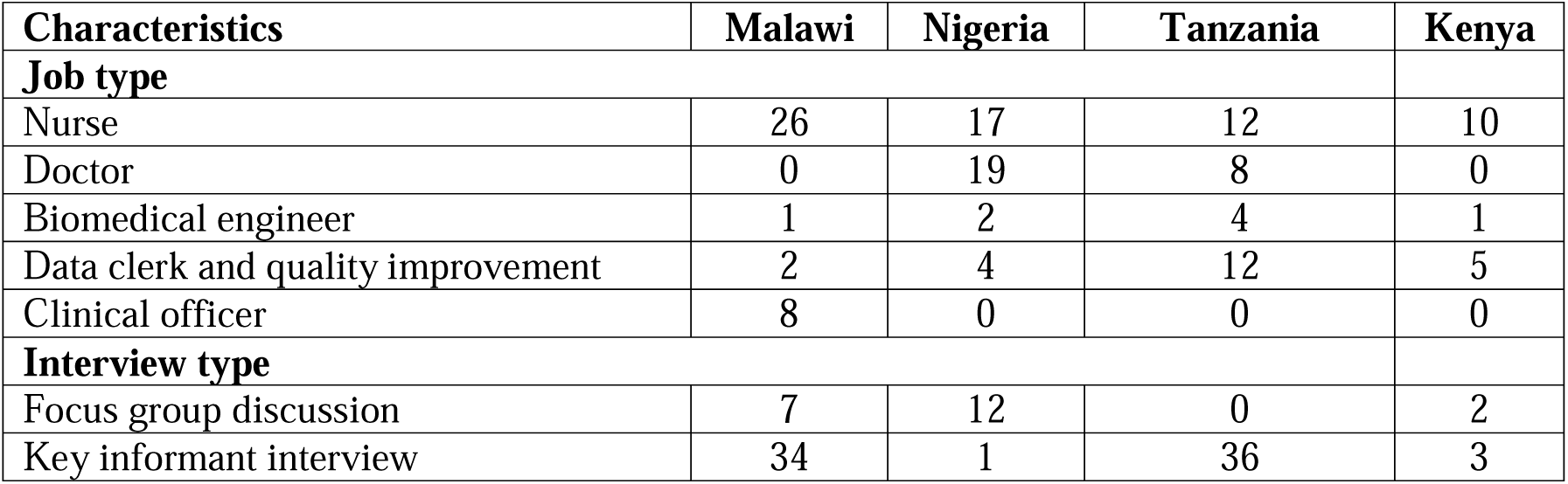
Participant Demographics.

### Strategies

A total of 114 distinct strategies were identified across several SSNC domains, including activities targeting post-discharge follow-up (3 strategy), financing (11 strategies), human resources (17 strategies), leadership (9 strategies), governance (6 strategies), data systems (11 strategies), quality improvement (QI) (13 strategies), infrastructure (7 strategies), equipment and commodities (15 strategies), infection prevention and control (IPC) (10 strategies), and linkage to maternal care which we expanded to family engagement (12 strategies). Table 2 provides a list of all strategies that were identified as well as the number of hospitals in each country that reported the activity. Table 3 shows which countries reported to use each strategy. Overall, 22 strategies were reportedly used by all four countries.

**Table 2.**
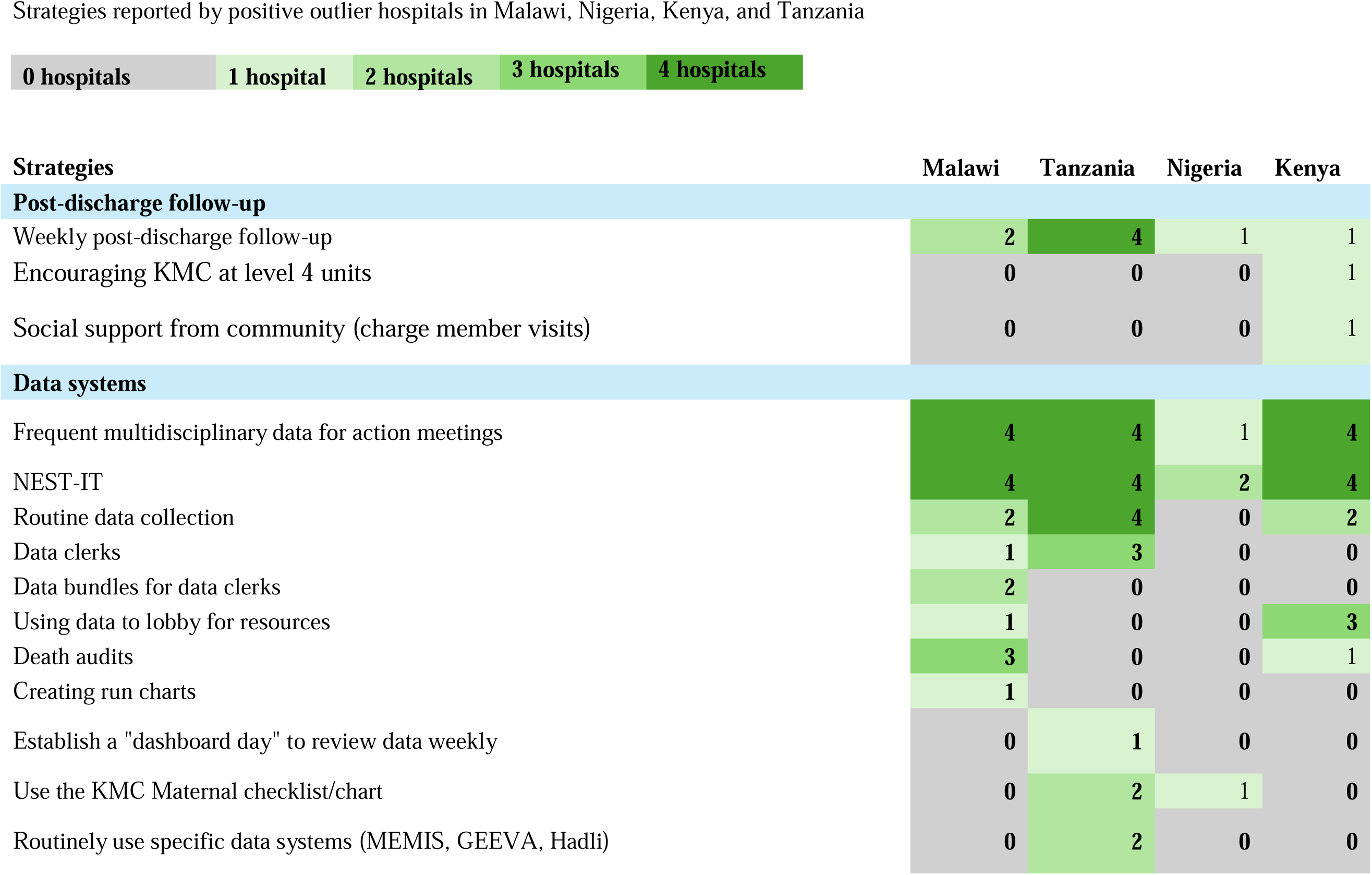
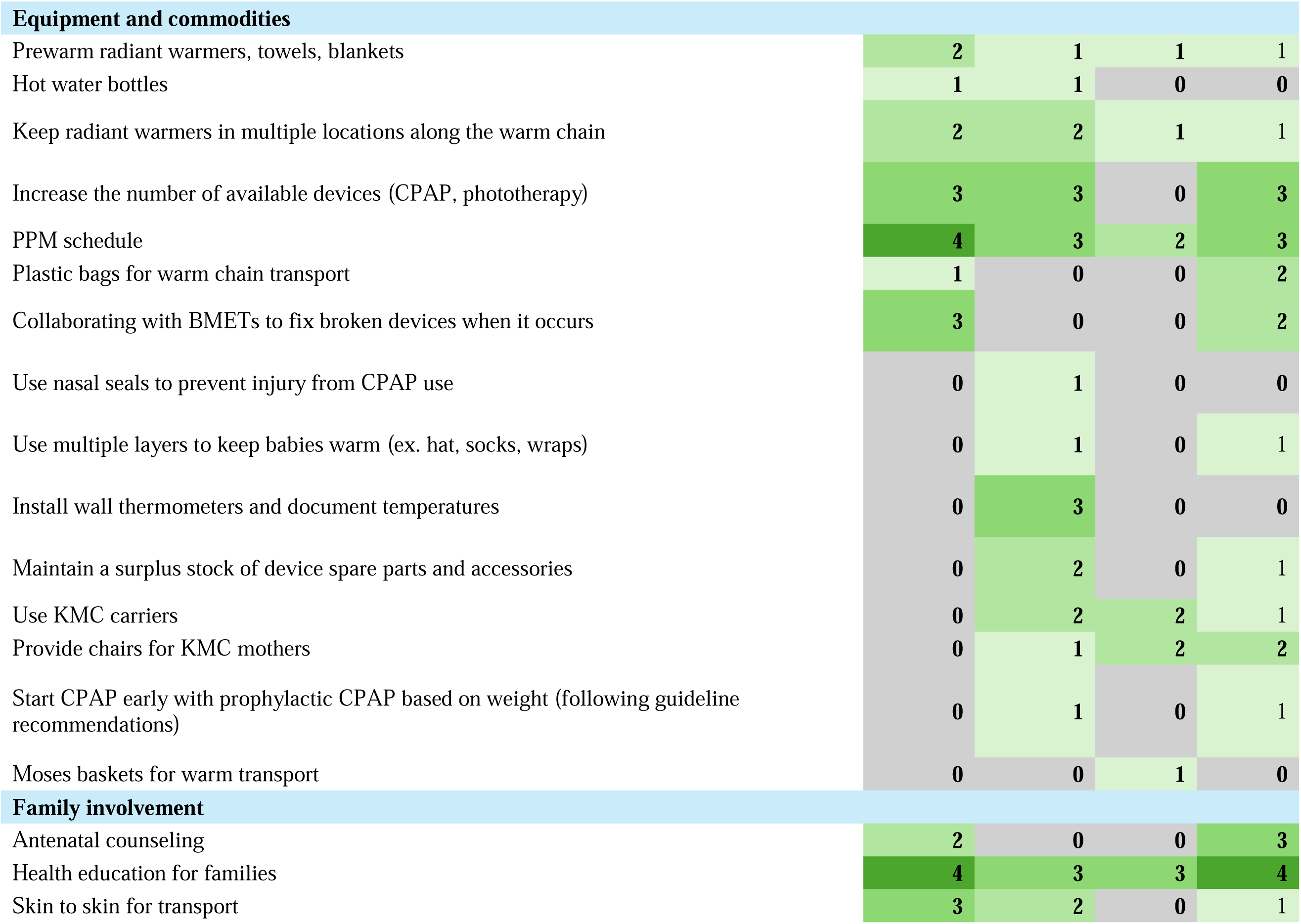
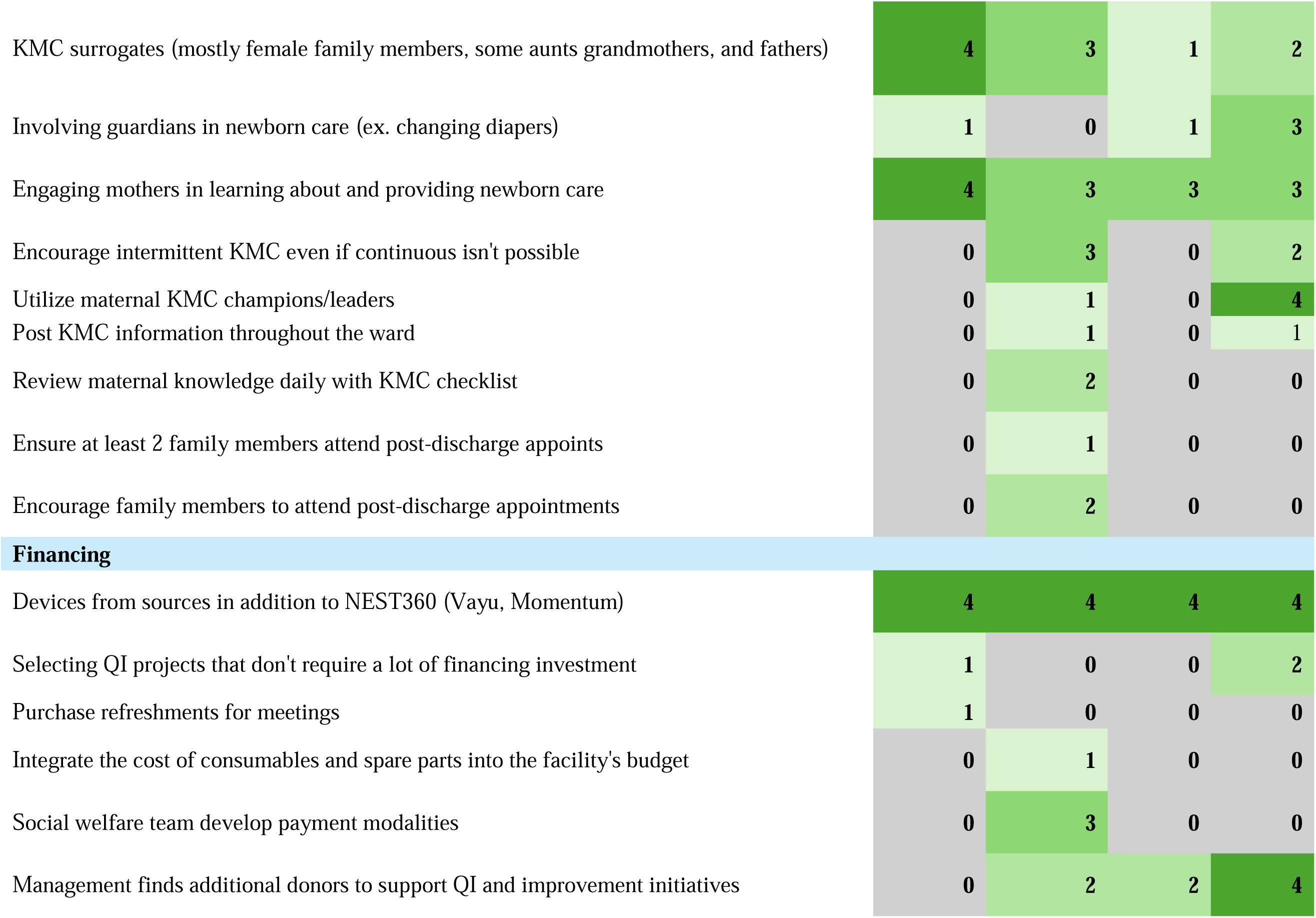
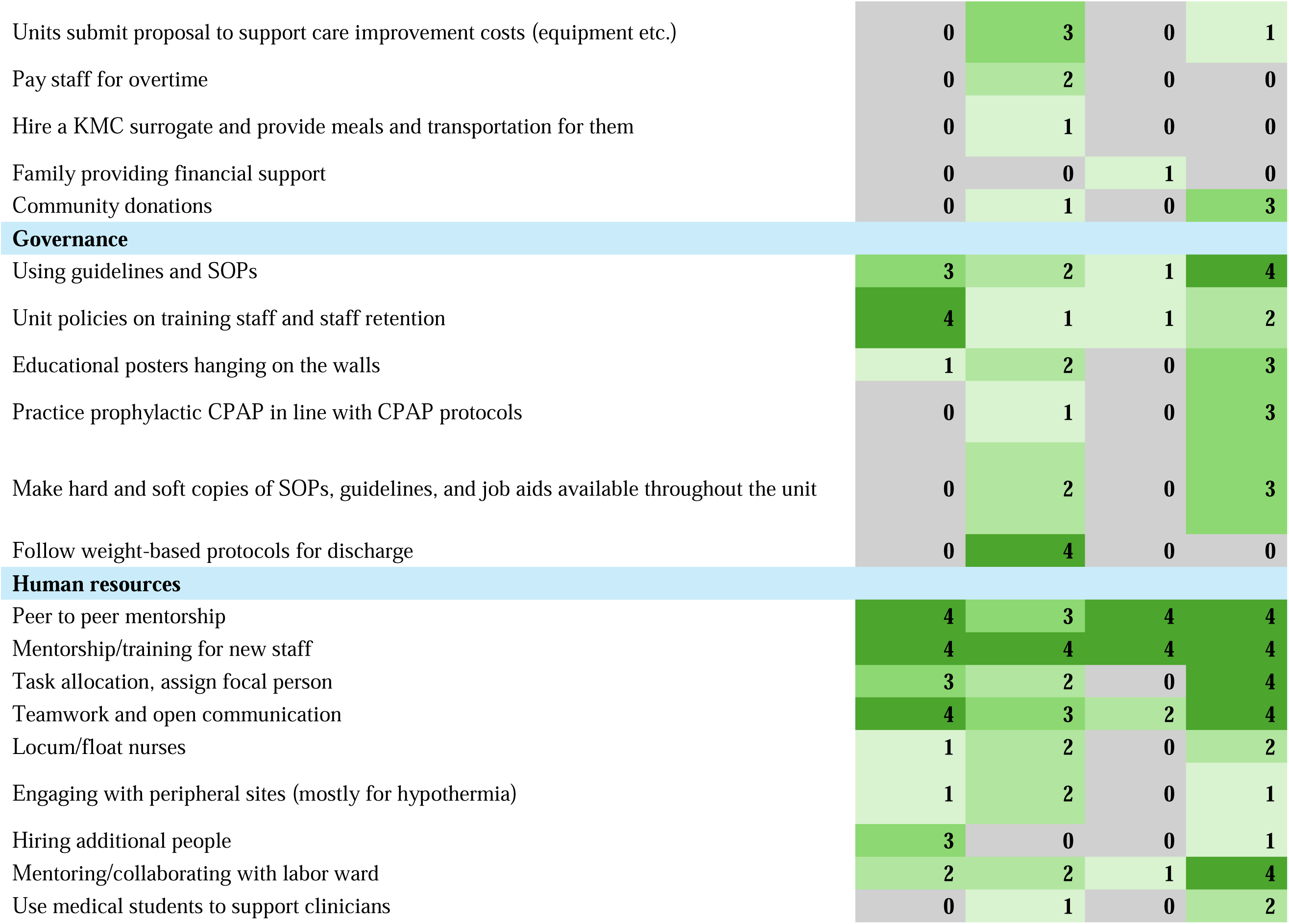
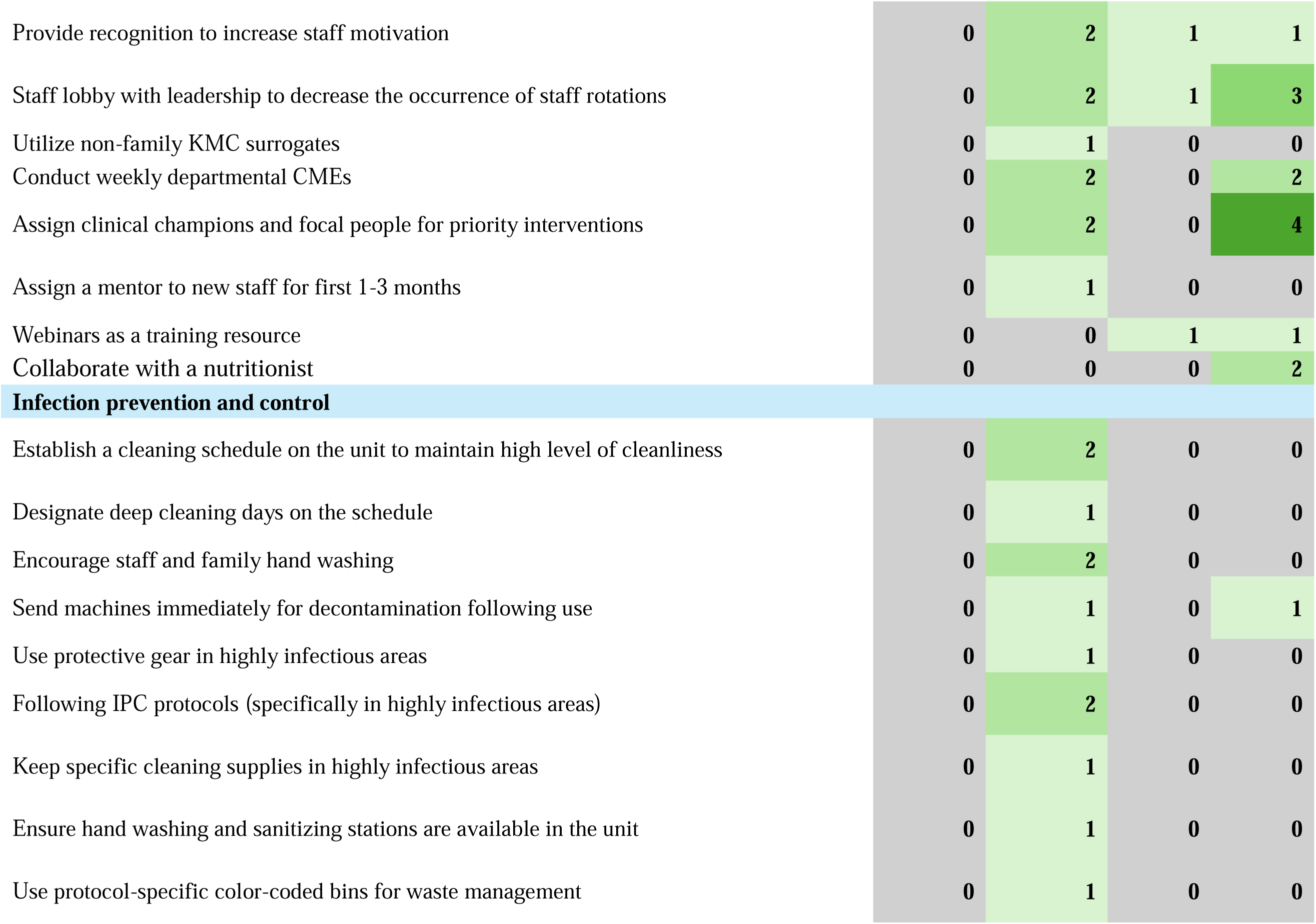
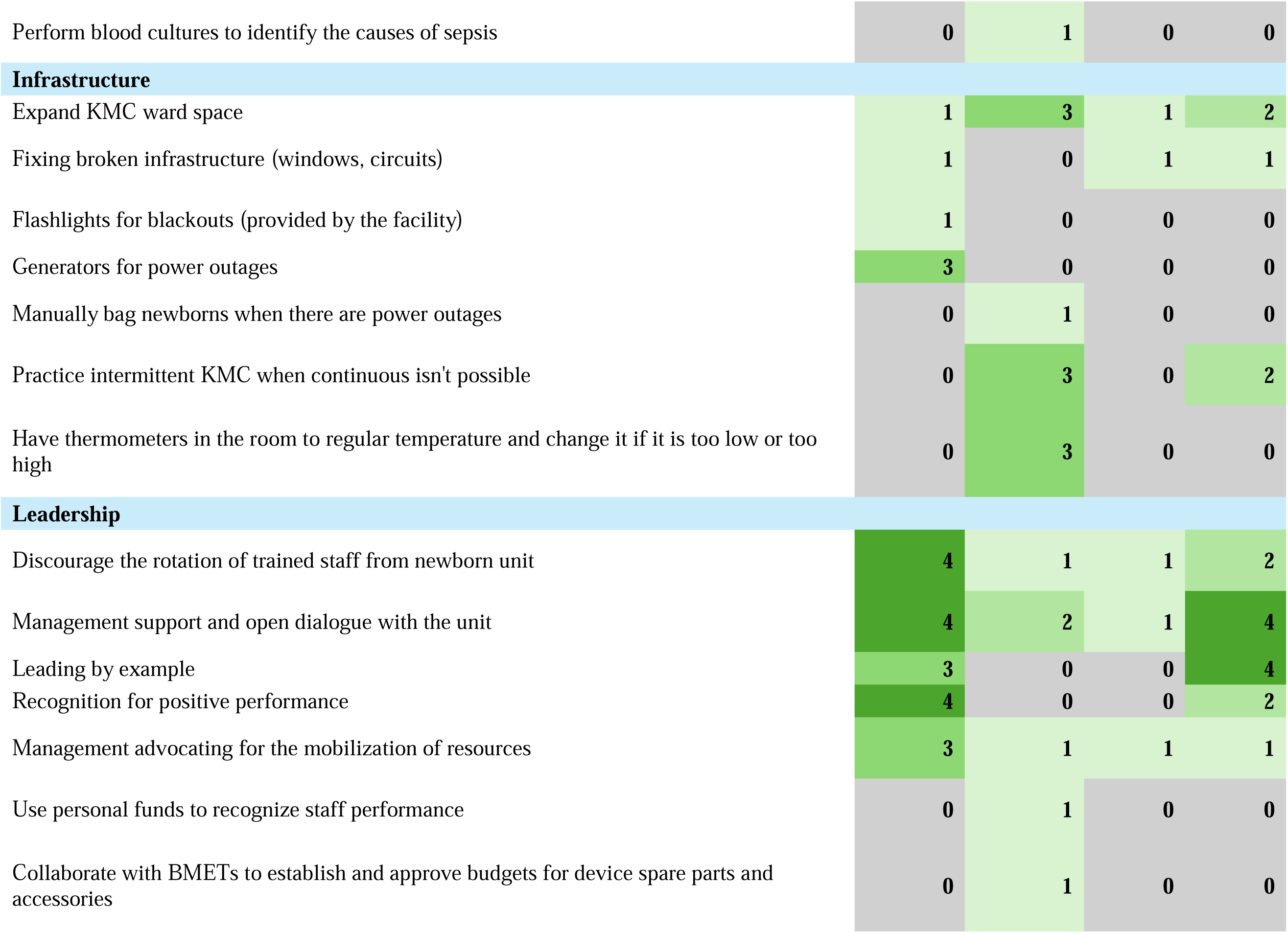
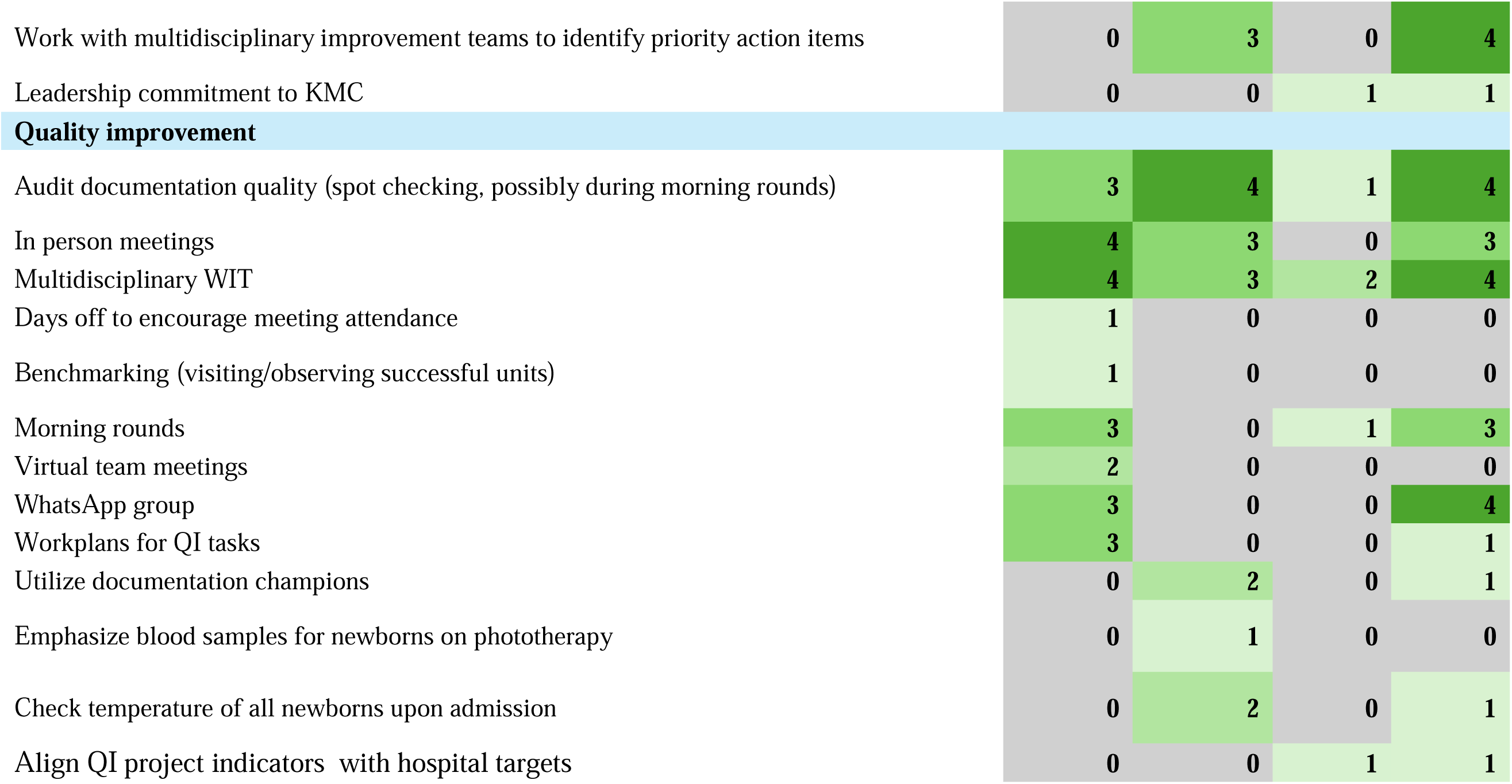
Frequency of strategies reported across positive outlier hospitals in Malawi, Nigeria, Kenya, and Tanzania.

**Table 3.**
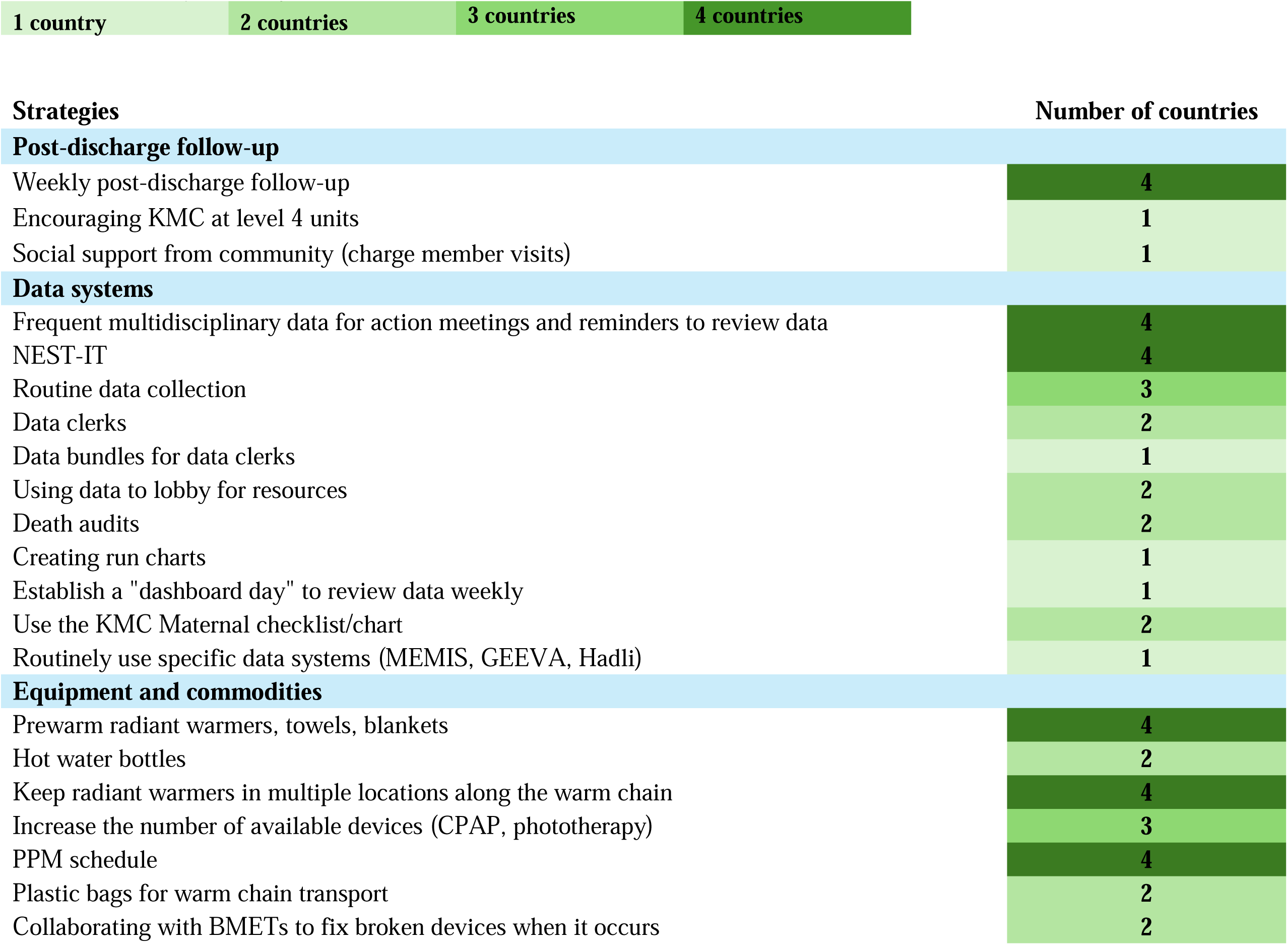
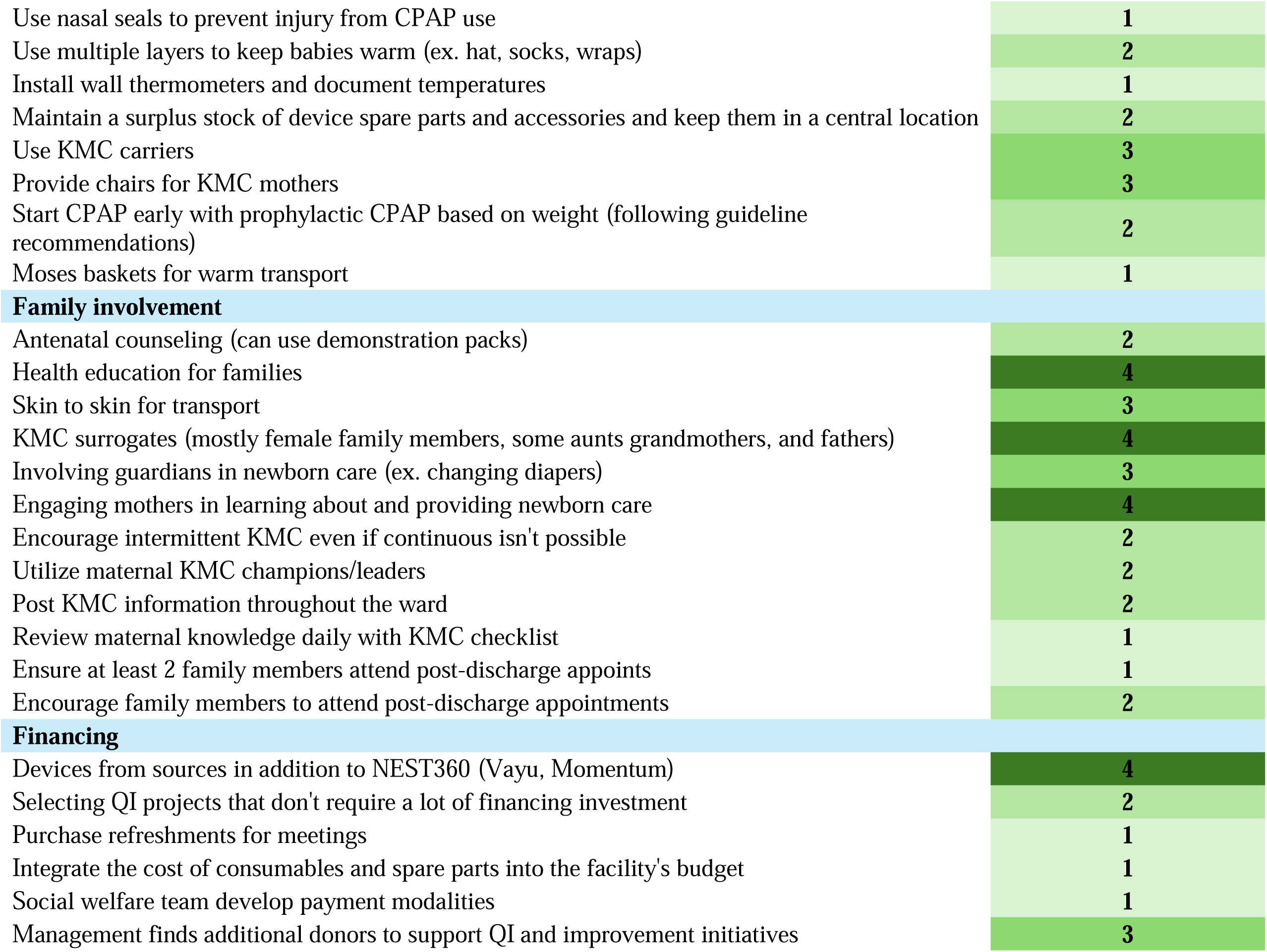
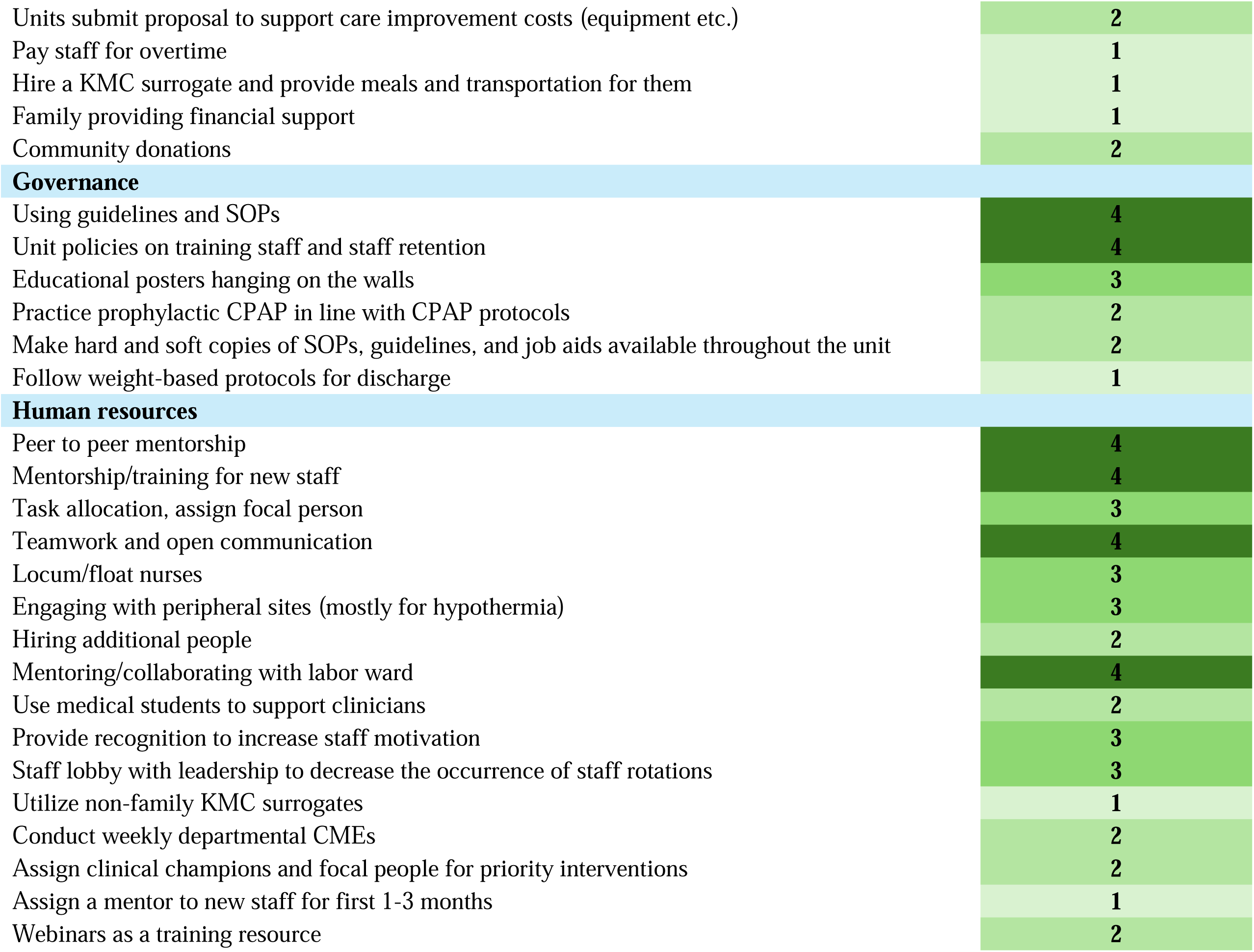
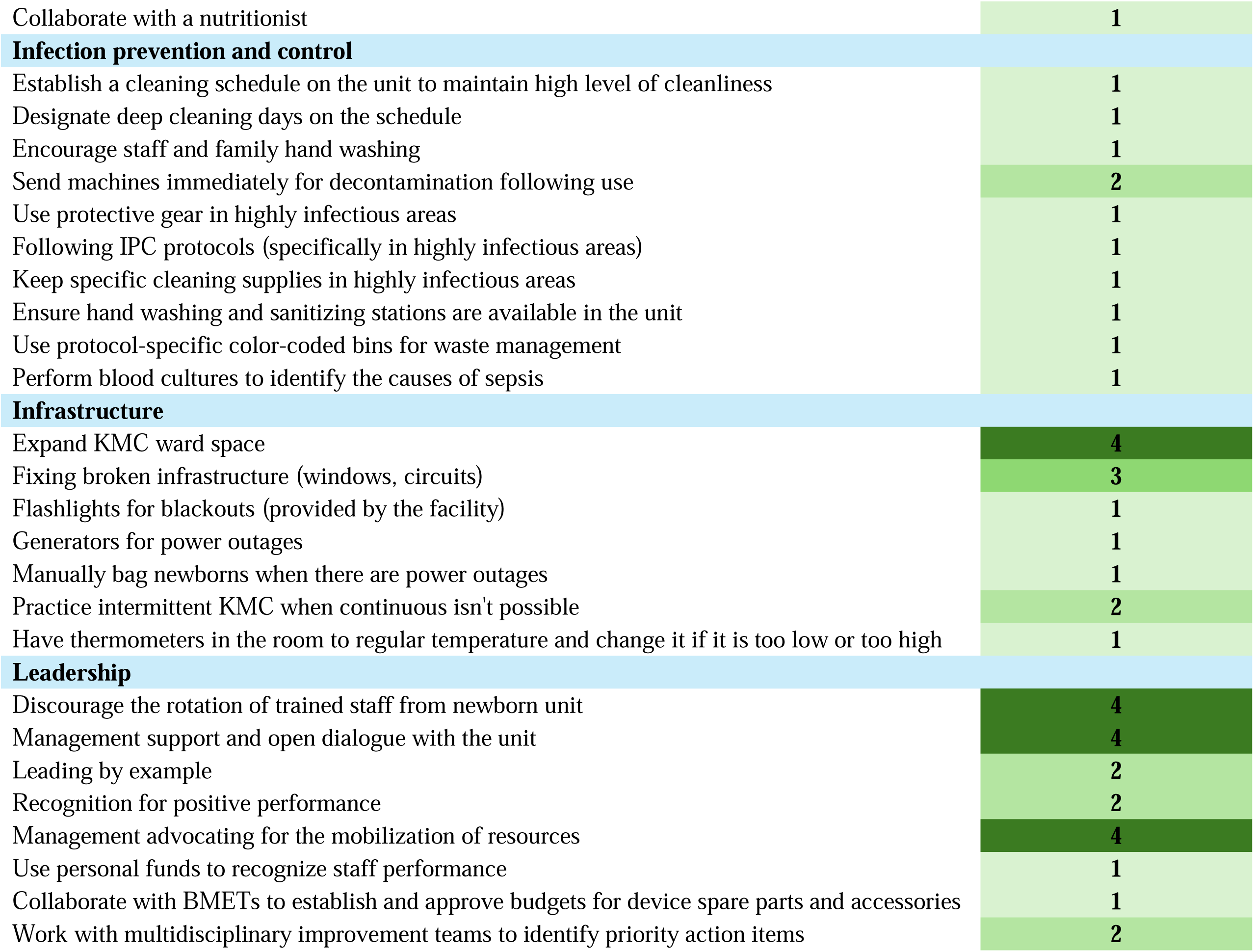
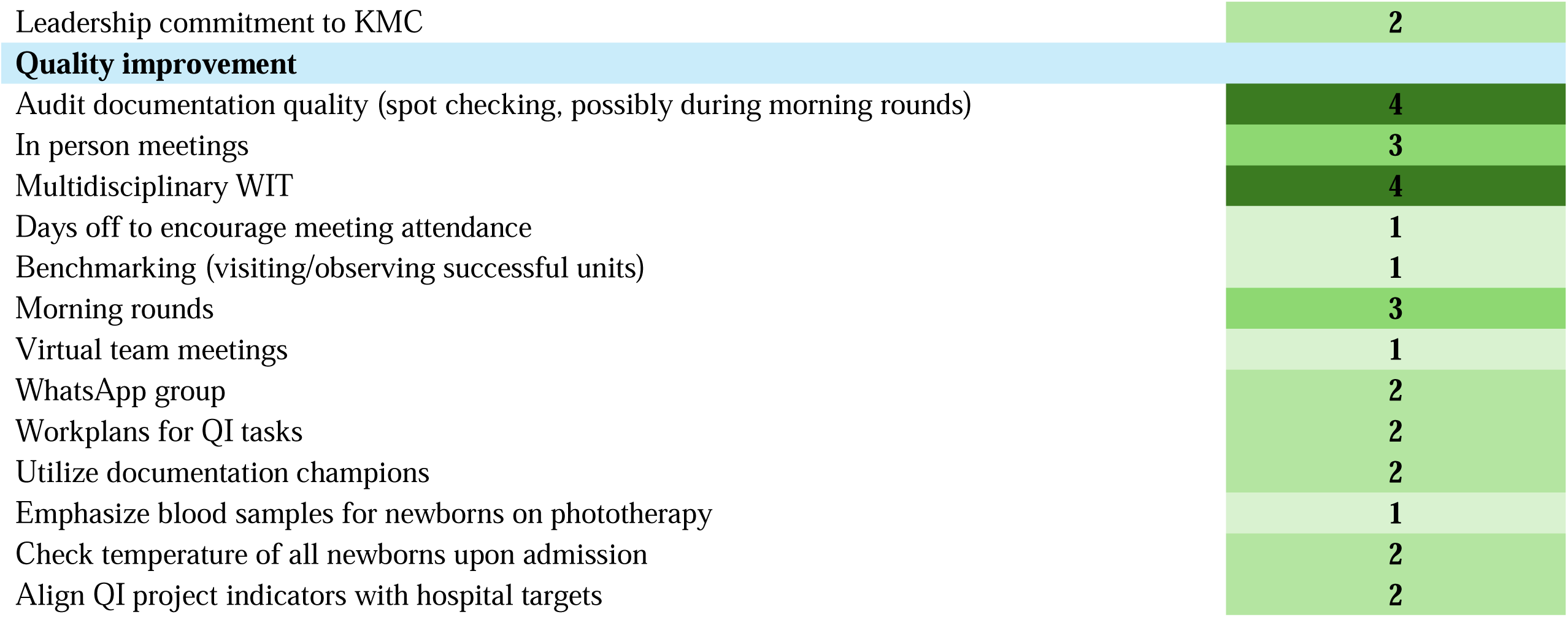
Strategies reported by positive outlier hospitals in Malawi, Nigeria, Kenya, and Tanzania.

These cross-country strategies included health education for families and active engagement of mothers in newborn care, which were consistently cited as critical for improving coverage for interventions such as KMC.

> *“We have been doing more awareness interventions for caregivers (i.e. mothers and relatives). The campaigns focused on the need and importance of KMC, especially in preterm or low birthweight babies to prevent hypothermia.”-Nigerian doctor*

Peer-to-peer mentorship and structured training for new staff were also universally identified as key strategies for ensuring high coverage rates of key indicators.

> *“Additionally, when new teams come in, they often spend one to three months with us, during which we assign a mentor to guide them on using the equipment. As a result, every new team member is well-prepared and knows how to operate the devices when they arrive.”* -Tanzanian Nurse

Finally, the formation of multidisciplinary ward improvement teams was also reported by all countries as a way to foster team engagement and identify effective change ideas for QI projects. This multidisciplinary collaboration was seen in ward improvement team membership of nurses, clinicians, biomeds, and unit leadership. Also, in some cases facilities had representation from multiple units including staff from labor and delivery as well as newborn units, which participants stated helped them to strengthen the continuity of care for newborns from delivery to admission in the newborn unit.

Furthermore, it was noted in some cases such as the one below, that mothers were included in the QI teams and process for identifying solutions.

> *“Firstly, we sat down as a team, both from the labor ward, and the neonatal unit, and we listed the problems that were causing the hypothermia for these babies. The we shared the tasks that this team should do…then working jointly this is what is happening now and keeping us going…from the labor ward, which is composed of members of the staff, the nurses, even the subordinate staff, and the hospital attendants from the labor ward, and from the nursery ward, there are also some nurses, we have clinicians, the data clerk, the hospital attendants as well, even the mothers themselves from both the labor and nursery ward”* -Malawian Nurse Midwife

While some strategies were commonly implemented across all countries, others were highly specific to local hospitals or national contexts. Hospitals in Malawi showed a strong emphasis on data-driven decision-making, frequently reporting the use of NEST-IT, the NEST360 program data dashboard, death audits, and multidisciplinary “data for action” meetings. Additionally, Malawi hospitals described strategies for internally sustaining QI activities, including selecting low-cost projects and purchasing refreshments for staff meetings. Tanzania, by contrast, described multiple strategies focused on IPC and infrastructure. For example, Tanzanian hospitals implemented strategies for IPC, such as designated deep-cleaning days and color-coded waste management systems. In Nigeria, while overall strategy reporting was less frequent, leadership engagement was a common theme. Furthermore, Nigerian hospitals described using personal funds to recognize staff, engaging in resource advocacy. Finally, Kenyan hospitals emphasized teamwork and interdisciplinary collaboration. While all countries discussed coordinating efforts with hospital biomedical technicians and communicating with labor ward staff, Kenya was the only country to report the inclusion of a nutritionist in the interdisciplinary newborn support team.

> *And then you see the importance of KMC, good nutrition, and working hand-in-hand with the nutritionist. We have a nutritionist who is allocated to take care of the KMC mothers.”* -Kenyan QI champion

Some innovative strategies emerged from individual hospitals. One Tanzanian hospital introduced “dashboard day,” dedicating a specific weekday for data review and performance tracking. Other unique approaches included the use of maternal KMC champions, involvement of family members in post-discharge care planning, and the hiring of KMC surrogates with financing support for meals and transport.

> *“We were fortunate to have a KMC champion, [redacted name], who volunteered to care [KMC] for abandoned babies.”* -Tanzanian Doctor

Malawi, Kenya, and Tanzania were active in preventative maintenance efforts for medical devices, such as establishing planned preventive maintenance (PPM) schedules and collaborating closely with biomedical engineers to ensure consistent device functionality.

> *“We make sure that all the equipment being used, the CPAP machines and concentrators, are all in good condition. We conduct regular PPM to make sure the outcome, say purity of oxygen being administered is within a good range. Another thing is that we make sure that all the machines are working properly, we try to move around for inspections according to the schedule. When a fault has been reported, we respond quickly.”* -Malawian Nursing and Midwife Officer

Conversely, some strategies were underreported or absent in specific contexts. For example, while Malawi, Kenya, and Tanzania frequently reported the use of a documentation champion or focal person, these strategies were not mentioned in Nigeria. However, Nigeria was the only country to reference webinars as a training tool.

Contextual factors between countries also played a role in the importance of change ideas. In three of the four countries (Kenya, Malawi, Tanzania), participants reported that their unit had used professional or family member surrogates to provide KMC to the newborn if the mother was unavailable. However, while Nigerian participants noted that KMC surrogates had been used in the past, they reported the practice was not effective. Participants stated that surrogates conflicted with cultural norms, as it was considered inappropriate for individuals other than the parents to provide skin-to-skin contact with the newborn, and for men and women to perform KMC in the same room due to the need to lower clothing for skin-to-skin positioning.

> *“In such case [where the mother is unavailable], it becomes more challenging because we don’t encourage fathers to perform KMC due to our constrained space, which may compromise the privacy of other mothers in the ward if a father is performing in.”* -Nigerian Doctor

Hospitals reported activities that targeted all the EBIs of interest (KMC, CPAP, phototherapy, and hypothermia prevention). Examples included using a weight-based protocol for CPAP initiation and transporting small newborns in plastic wraps from the labor to neonatal ward as a hypothermia prevention strategy. However, while some strategies were closely linked to specific indicators, others were more general and promoted positive outcomes for more than one indicator. For example, some of the change ideas promoting KMC ultimately also supported improvements in hypothermia prevention. These change ideas included things such as transporting newborns in a skin-to-skin position or using family members, such as fathers and grandmothers, to act as KMC surrogates if the mother was unavailable. Another example of change ideas supporting more than one EBI was the use of PPM to ensure devices remained functional which supported the utilization of both CPAP and phototherapy machines.

## 4. Discussion

This qualitative study identified 114 distinct strategies that NEST360-supported positive outlier hospitals used to improve or maintain a high level of SSNC in neonatal units. The broad list of activities underscores the importance of exploring contextual differences and barriers that can be present between hospitals, countries, or regions within multi-site programs. This allows implementers to identify needed adaptations that support targeted implementation, even if all sites in the program formally receive the same core package of interventions, such as NEST360’s technology, training, data systems, and QI support. The discrete activities that were used at each site to implement the NEST360 package varied within a country and across multiple countries, as seen by the varied coverage rates and reported activities across sites. These differences in strategy selection and implementation could be driven by multiple facility and country level differences including those impacting leadership, infrastructure, policy, and employee capacity. By identifying high performing “positive outlier” hospitals, we were able to uncover strategies that high performing hospitals were using to improve and maintain coverage for interventions such as KMC, CPAP, hypothermia prevention, and phototherapy. These findings provide actionable insights that can guide slower-performing NEST360 hospitals, future NEST360-supported countries and hospitals, as well hospitals outside the alliance, toward more effective adoption of SSNC practices.

However, it is important to note that just because a participant at a specific facility did not report a strategy this does not mean that it could not have been happening there. Prior to introducing new strategies into a site, the implementer should investigate if those activities are already occurring in any capacity.

Our findings echo a growing body of implementation science research demonstrating that variability in care outcomes is common, even when hospitals receive the same interventions and resources (24, 25). The observed heterogeneity in implementation success underscores the need for adaptive, locally tailored strategies supported by cross-country learning. The qualitative descriptive approach used here facilitated a deeper understanding of what is truly occurring at high performing hospitals. This level of insight is especially valuable for program scale-up and sustainability planning, as it goes beyond measuring outcomes to understanding the mechanisms behind them. For example, not all implementation strategies are universally applicable across contexts, a pattern that emerged clearly across the four countries included in this study. For example, we observed varying levels of receptiveness to KMC surrogates in Tanzania and Nigeria. This aligns with existing literature, which has similarly reported that the success of KMC surrogate use depends on the specific implementation context (26, 27). This illustrates the importance of aligning implementation strategies with local cultural expectations and care practices.

Many of these successful strategies worked not only because of their technical merit, but also because they addressed deeper organizational and behavioral mechanisms, such as motivation, accountability, and trust, which influence care delivery (28, 29). For example, routine, multidisciplinary data review meetings using the NEST-IT dashboard, provided a space where teams could regularly review their facility’s performance data, which can foster a culture of transparency and collective responsibility. These practices encouraged reflection on progress, promoted teamwork, and reinforced staff ownership of progress.

Other strategies, such as peer-to-peer mentorship or role rotation in clinical and support duties, were identified as useful across multiple settings. These activities may have worked by strengthening internal capacity and reducing dependency on a small number of trained individuals who could be rotated to other units. These approaches can also create informal networks of accountability, where staff feel comfortable asking others for support.

Additionally, visible commitment from leadership, such as allocating resources, recognizing staff contributions, or personally participating in improvement activities like QI meetings, served as a powerful motivator to staff that their work was important (30–32). Taken together, these examples illustrate that beyond technical fixes, like fixing broken doors and windows, strategies that nurture a sense of agency and teamwork may be especially effective in sustaining high-quality care in low-resource settings.

### Comparison to existing literature

While previous studies have documented barriers to implementation or described QI efforts within individual countries or regions, few have systematically mapped facility-level strategies for SSNC in higher performing sites across multiple countries to identify generalizable patterns (33). This study adds to the literature by going beyond surface-level descriptions to examine how and why specific strategies were implemented based on the barriers in their unique context. In doing so, it adds depth to prior work on neonatal health in low-resource settings by centering the perspectives of frontline staff and highlighting the locally tailored solutions they developed.

Moreover, the cross-country, multi-site design of this study allowed for the identification of convergent strategies that proved successful across diverse contexts, as well as culturally or structurally specific adaptations. These insights align with recent calls in global health to move from “what works” to understanding “what works, for whom, and under what circumstances” (34). By documenting both the diversity and similarities of change ideas, this study provides an empirical foundation for designing more adaptive and context-responsive implementation strategies in neonatal care.

Furthermore, this research highlights how successful QI efforts, though deeply rooted in local context, can yield lessons of broader relevance. Facility teams leveraged a range of context-specific strategies, from task-shifting to strengthening intern-unit collaboration. By analyzing these approaches across countries, we identified common themes and extracted actionable lessons that can be adapted to improve care in other low-resource settings. This process of leveraging facility-level innovations to identify generalizable lessons that can be applied across diverse contexts aligns with current calls from implementers and researchers to leverage tools and strengths from QI and implementation science (35–39). Implementers should consider investing in mechanisms for intra-country or cross-country learning to elevate locally identified successful strategies that could be applicable to a broader audience. These mechanisms could be similar to formal qualitative inquiry, such as an exemplar study (40), or general formats like learning collaboratives.

### Future Directions

Next steps include quantitatively exploring the correlation between specific facility-level change ideas and improvements in care quality. Understanding these relationships will provide a stronger evidence base for recommending strategies with the greatest impact. Additionally, integrating this research with routine monitoring and evaluation systems will support ongoing learning and adaptation within NEST360 and similar implementation programs. Furthermore, during the data collection process, the Nigerian team identified negative outlier facilities as well as their positive outlier, so the team will explore the frequency of reported strategies between the two types of outliers, which will help to uncover additional details related to the influence of specific strategies on coverage rates and care delivery. Based on findings this negative outlier analysis may be replicated in Kenya and Malawi as well.

### Limitations

This study has several limitations. First, we were unable to conduct a one-to-one comparison of which specific strategies directly contributed to changes in clinical coverage. Most strategies were implemented concurrently or as a part of a broader QI package, limiting our ability to isolate the impact of individual activities. Second, while we aimed for consistency across sites, there were variations in data collection methods (e.g., FGDs vs. KIIs) and participant types across countries. These differences may have influenced the depth and scope of insights captured. Also, our reliance on facility staff reporting may have missed undocumented or informal practices, and simply because a facility did not report an activity does not mean that it was not occurring there. Although our coding approach allowed for inductive theme development, the analysis was primarily guided by a deductive, a priori framework. This may have introduced some interpretive bias, potentially constraining the emergence of unanticipated themes.

Additionally, there is a risk of social desirability bias in participant responses, as staff may have emphasized successful strategies or downplayed challenges to make their hospitals look better. The study focused on positive outliers and did not include negative outliers, so findings may not fully represent challenges faced by lower-performing hospitals. Finally, this analysis selected participating sites based on care coverage and did not measure the quality of the implementation of EBIs.

## 5. Conclusions

This study highlights the critical role of facility-level strategies in shaping the implementation and coverage of essential newborn interventions within a multi-country program. Despite receiving the same core support from NEST360, hospitals varied in their ability to consistently deliver high-quality care, underscoring the influence of local adaptation, leadership, and context. By systematically examining the strategies employed by high-performing hospitals, we identified actionable, often low-cost approaches that can inform broader implementation efforts in similar low-resource settings. These findings reinforce the value of adaptive QI, cross-country learning, and the intentional documentation of contextualized solutions. Embedding such practices into routine health system strengthening efforts may accelerate progress toward reducing neonatal mortality and improving newborn outcomes globally.

## Supporting information

SRQR checklist

## Data Availability

Data are available upon reasonable request. The data set used during the current study is fully owned by the hospitals that provided the information. Data that could be linked to a particular facility will not be shared outside of NEST. Aggregated data (multicountry and program level) may be made available at the discretion of the NEST360 Steering Committee. Requests for data access can be made on the NEST360 website in the Request for Public Use of NEST360 Data section. Many of the tools and educational materials mentioned in this manuscript are already available on the NEST360 website in the Resources section. Requests for additional tools, materials, and learning are available from the NEST360 Steering Committee upon reasonable request.

## 6. List of abbreviations

(EBIs): Evidence-based interventions
(KMC): Kangaroo mother care
(CPAP): Continuous positive airway pressure
(NEST360): Newborn Essential Solutions and Technologies
(QI): Quality improvement
(UNICEF): United Nations Children’s Fund
(SSNC): Small and sick newborn care
(SRQR): Standards for Reporting Qualitative Research
(BMETs): Biomedical engineering technicians
(KIIs): Key informant interviews
(FGDs): Focus group discussions
(IPC): Infection prevention and control
(PPM): Planned preventive maintenance

## 7. Declarations

### Ethics

Ethical approval for this study was conducted by London School of Hygiene and Tropical Medicine IRB (21892), and the national health research ethics committee in Malawi (NHSRC 1180), Kenya (Kenya Medical Research Institute: KEMI/SERU/CGMR-C/229/4203, Maseno University Ethics Review Committee: MSU/DRPI/MUERC/00810/19), Tanzania (Ifakara Health Institute: IHI/IRB/No:01-2020, National Institute for Medical Research: NIMR/HQ/R.8a/Vol.IX/3405), and Nigeria (Lagos University Teaching Hospital Health Research Ethics Committee: ADM/DCST/HREC/APP/3487, University of Ibadan/University College Hospital/Ethics Committee: NHREC/05/01/2008a).. The study was conducted with the ethical principles outlined in the Declaration of Helsinki. All participants were informed about the purpose of the study, assured of confidentiality, and provided verbal consent prior to participation. Participation was voluntary, and respondents could withdraw at any time without consequence.

### Consent for publication

Not applicable.

### Author contributions

All authors made substantial contributions to the development of the manuscript. KD, CB, LH, LM, JL, ZO, and RRK conceptualized the study. KD coordinated the overall project and led the drafting of the manuscript. IO, HM, JB, SN, LM lead country-level data collection and data analysis with support from KD. LM, MJ, CE, KK, RT, DS, NR contributed to critical revisions and final approval of the manuscript, and all authors agreed to be accountable for all aspects of the work.

## Acknowledgements

First, and most importantly, we thank the newborns and their mothers whose data are the heart of NEST360. We also thank those involved in the day-to-day program activities, including the clinicians, biomeds, and managers at the individual hospitals. Many thanks to the relevant administrative staff for their support. We are also very grateful to the fellow researchers and editors who peer-reviewed this paper and for the input from the managing editors at *BMC Health Services Research* and within NEST360. We acknowledge that this work has not been published or presented before.

Additionally, we want to acknowledge the facility staff who enabled this work to be possible and are identified as group authorship, and include the following individuals, Veronica Ongwae, Nancy Mburu, Jackline Otieno, Jacinta Moki, Susan Nangila, Dina Khaemba, Maureen Natembeya, Victorina Makona, Stanley Limo, Doreen Okello, and Nicholas Kipketer from Kenya, Jenipher Kamera, and Alice Khonje from Malawi, Joseph G. Kimaro, Bryceson L. Kiwelu, Martha Nkony, Rehema Marando, Mark B. K. Charles, Mary Machemba, Grace Mhando, Bernadetha Shine, Salha Omary, Tumaini Bayasabe, Gilson Mwampegete, Julieth Bachibila, Secilia Ndaki, Stella Mmasi, Mtagi Kibatara, Felix Bundala, Martha Nkony, Rehema Malando, and Mary Charles from Tanzania, Iretiola Bamikeolu Fajolu, Beatrice Nkolika Ezenwa, Olaolu Aziza Moronkola, Olukemi Oluwatoyin Tongo, Michael Abel Alao, Olutekunbi Olanike Abosede, Ayodele Abiona, Titilayo Kayode-Alabi, Oma Nnenna Amadi, and Marilyn Ene Okpe from Nigeria.

