## Supplemental Table 1 for "Uncovering and Understanding Success: A Qualitative Study of High-Performing Hospitals for Small and Sick Newborn Care in Four countries in Africa"

**Supplemental Table 1.** Breakdown of the different indicators each positive outlier met

|  | **Rapid progressor** | | | | **Sustainer** | | | |
| --- | --- | --- | --- | --- | --- | --- | --- | --- |
| **Outlier** | **CPAP** | **KMC** | **HP** | **PJ** | **CPAP** | **KMC** | **HP** | **PJ** |
| Kenya 1 |  |  |  |  |  |  |  |  |
| Kenya 2 |  |  |  |  |  |  |  |  |
| Kenya 3 | *Selected by the country team based on contextual insight, and knowledge of current quality improvement activities* | | | | | | | |
| Kenya 4 | *Selected by the country team based on contextual insight, and knowledge of current quality improvement activities* | | | | | | | |
| Malawi 1 |  |  |  |  |  |  |  |  |
| Malawi 2 |  |  |  |  |  |  |  |  |
| Malawi 3 |  |  |  |  |  |  |  |  |
| Malawi 4 |  |  |  |  |  |  |  |  |
| Nigeria 1 |  |  |  |  |  |  |  |  |
| Nigeria 2 |  |  |  |  |  |  |  |  |
| Nigeria 3 |  |  |  |  |  |  |  |  |
| Nigeria 4 | *Selected by the country team based on contextual insight, and knowledge of current quality improvement activities* | | | | | | | |
| Tanzania 1 |  |  |  |  |  |  |  |  |
| Tanzania 2 |  |  |  |  |  |  |  |  |
| Tanzania 3 |  |  |  |  |  |  |  |  |
| Tanzania 4 |  |  |  |  |  |  |  |  |

*Note.* Continuous positive airway pressure (CPAP); kangaroo mother care (KMC); hypothermia prevention (HP); phototherapy for jaundice (PJ)
